# Self-cleaning menstrual cups with plant-based biodegradable superabsorbent fibrous tablets for hygienic and sustainable Period Care

**DOI:** 10.1101/2025.05.08.25327253

**Authors:** Shaghayegh Moghimikandelousi, Fereshteh Bayat, Lubna Najm, Rida A. Malik, Aishwarya Bhavan, Farbod Azaripour Masooleh, Leisa Hirtz, Jeffrey I. Weitz, Zeinab Hosseinidoust, Tohid F. Didar

**Affiliations:** School of Biomedical Engineering, McMaster University, 1280 Main Street West, Hamilton, ON, Canada; Department of Mechanical Engineering, McMaster University, 1280 Main Street West, Hamilton, ON, Canada; Thrombosis & Atherosclerosis Research Institute and Hamilton Health Sciences, 237 Barton Street East, Hamilton, Ontario L8L 2X2, Canada; Department of Medicine, McMaster University, 1280 Main Street West, Hamilton, ON, Canada; Department of Biochemistry and Biomedical Sciences, McMaster University, 1280 Main Street West, Hamilton, ON, Canada; Department of Life Sciences, McMaster University, 1280 Main Street West, Hamilton, ON, Canada; Women’s Global Health Innovations, B21 - 175 Longwood Road South, Hamilton, ON, L8P 0A1 Canada; Department of Chemical Engineering, McMaster University, 1280 Main Street West, Hamilton, ON, Canada; Farncombe Family Digestive Health Research Institute, McMaster University, Hamilton, ON, Canada; Michael DeGroote Institute for Infectious Disease Research, McMaster University, Hamilton, Ontario, Canada

**Keywords:** Eco-friendly menstruation, menstrual cup, superabsorbent fibrous tablets, Feminine hygiene, Self-cleaning

## Abstract

Menstrual cups are a sustainable and reusable alternative to pads and tampons, offering reduced waste and lower risks of allergies, infections, and toxic shock syndrome. However, their widespread adoption and long-term use are hindered by hygiene concerns, specifically biofouling and the need for messy removal. We report a self-cleaning menstrual cup with a biocompatible silicone oil-infused coating combined with a biodegradable, plant-based tablet composed of superabsorbent fibers. The absorbent fibers were designed with crosslinked alginate and optimized to achieve high absorption capacities (8 mL water or 15 mL blood per g fiber) within 8 hrs. At the same time, the lubricant-infused cups prolonged clotting times and exhibited self-cleaning properties, with little blood detected on the treated cups and a significant reduction in bacterial adhesion/biofilm formation. The fibrous tablet retained blood in an amount equivalent to the full capacity of the menstrual cup, eliminating spillage during removal and facilitating easy emptying. By removing barriers to comfort and convenience, our design and developed materials present a simple and effective advancement in feminine hygiene products that tip the scale in favour of reusable products and advance global environmental stewardship.

---

Menstruation is a physiological experience shared by 1.8 billion humans every month, resulting from cyclical hormonal changes.^1–3^ Menstruation typically begins during puberty and on average ends with menopause.^3^ Despite nearly a quarter of the world’s population menstruating, menstrual hygiene products have remained unchanged for decades, with innovations hampered by stigma surrounding menstruation, as well as institutionalized misogyny still lurking in many corners of academia and industry.^4–9^ The need for innovation in the field is especially critical for women in low- and middle-income communities, where lack of appropriate menstrual products not only impacts their physical and mental health but their access to education and degree of involvement in communities.^3–8^

Tampons and pads are the most common solutions for absorbing menstrual blood. These have significant drawbacks, including a less-than-desirable environmental footprint and a potential for adverse health effects. Every year, 28,114 tonnes of waste are created from menstrual products in the UK alone, which extrapolates to millions of tonnes worldwide.^10^ Most pads and tampons are single-use and not biodegradable and may take 500-800 years to disintegrate in the environment.^11^ Additionally, the use of pads has been documented to cause rash, irritation, and allergy, and tampons have been infamously associated with toxic shock syndrome (TSS), a potentially fatal condition caused by bacterial infection.^12^ Reusable menstrual cups are gaining traction in the market as a more environmental and budget-friendly alternative to single-use pads and tampons, with one cup lasting 5-10 years.^6^ Cups have a 3-6 times higher capacity for holding menstrual blood compared to tampons (1-2 ounces vs a third of an ounce) and have not been associated with major health concerns.^13^ Although posed to be a game changer in the field, the widespread adoption of menstrual cups is hampered by the need for continual cleaning and a messy removal process with high chance of spilling.^13^

To address this issue, we envisioned a highly absorbent tablet to be placed on the surface of a menstrual cup, and treated to repel biosorption, resulting in a more hygienic experience. We synthesized the fibers from alginate hydrogels using a wet spinning setup followed by air-drying to obtain a dry porous superabsorbent flexible material (**Figure 1a**). Alginate is a natural superabsorbent polymer derived from brown algae that is biocompatible and non-immunogenic.^14–18^ It gains its superabsorbent properties from sodium acetate (-COONa) and hydroxyl (-OH) groups and can be crosslinked with divalent cations such as Ca^2+^, Mg^2+^, and Zn^2+^ to form bulk hydrogels and fibrous materials.^14,19^ Our choice of alginate crosslinked with calcium cations was reinforced by the documented potential to initiate clotting and further improve the blood-holding capacity of the material.^20–22^

**Figure 1.**
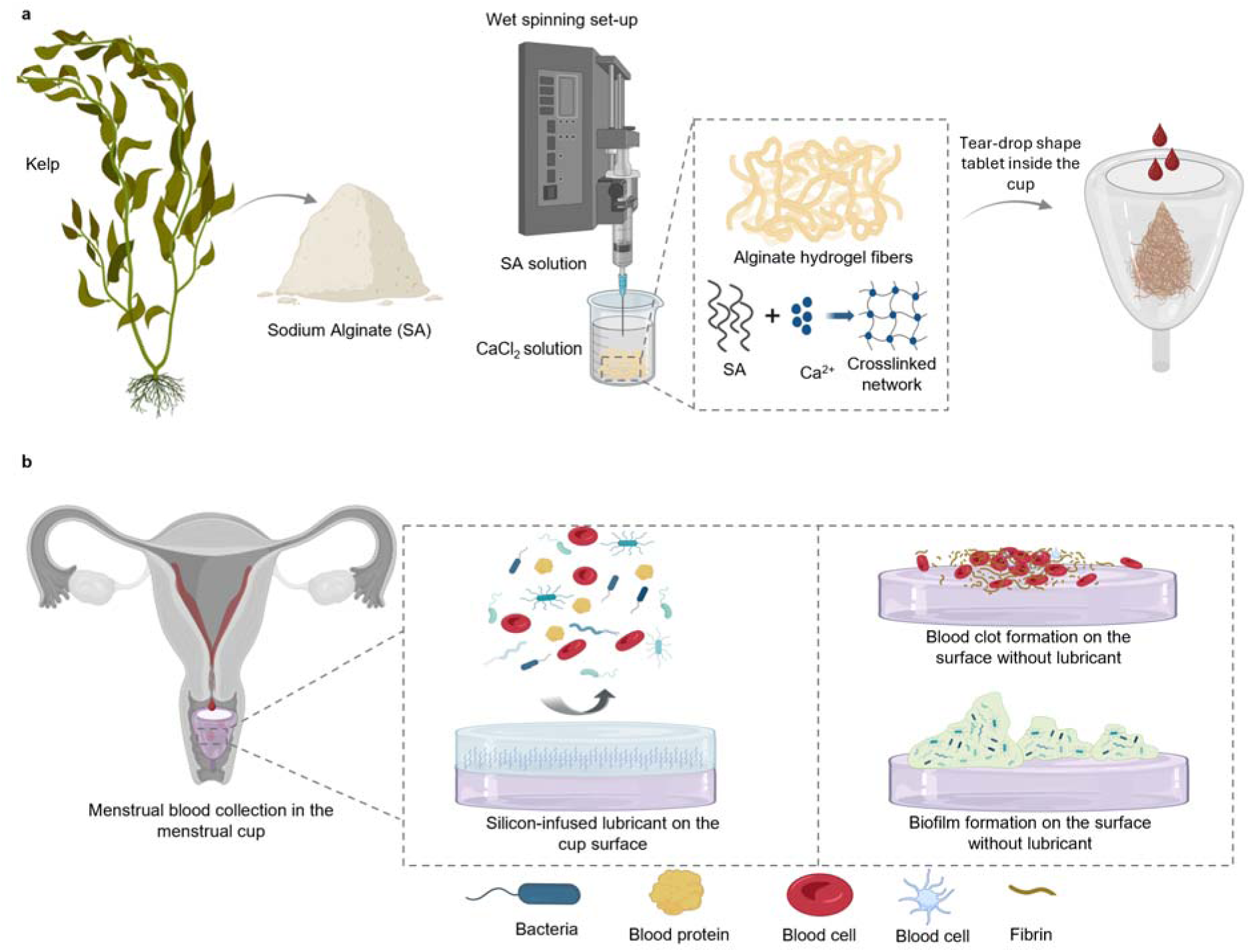
(a) Schematic representation of the alginate fiber production process using a wet-spinning setup and their function as a superabsorbent material for collecting menstrual fluid inside a reusable cup. (b) Biofilm and blood clot formation on the cup surface in the absence of lubricant.

Lubricant-infused surfaces (LISs) are a class of engineered coatings that create a stable, slippery liquid interface capable of resisting unwanted adhesion by biological substances.^23,24^ More recently, LIS coatings have emerged as an effective method for preventing nonspecific adhesion and biofilm formation in the various field such as biosensors^25,26^, medical devices like vascular graft^24,27^, catheter and implants^22,23,28–32^. The mechanism of LISs relies on the infusion of a chemically compatible lubricant into a porous or textured solid, forming a liquid layer that repels blood, bacterial cells, and biofilms by minimizing contact with the underlying substrate. In addition, this omniphobic nature reduces protein adsorption and cellular adhesion, thereby preventing clot formation and bacterial colonization.^23,24,28–35^ For this objective, the cup selected was lubricant-infused coating with silicon oil, which reduced biofilm formation and clot adherence, making for a self-cleaning cup (**Figure 1b**).^27,32,36^ The cup and absorbent fibers/tablet were characterized mechanically as well in human blood to monitor thrombin generation and clot formation and adherence. We further demonstrated the performance of the cup-tablet system by showing bacterial repellency and biofilm prevention.

## Result & Discussion

### Menstrual cup material characterization

We started by characterizing three different commercially available menstrual cups, cup #1, cup #2, and cup #3 (**Figure 2a**). While all commercially available cups are fabricated from medical grade silicone, their capacity ranges from 15 to 30 mL. Cup #3 was infused with silicon oil during manufacturing, unlike the other two samples. As shown in **Figure 2b**, the lubricant-infusion of cup #3 during manufacturing did not significantly affect the static contact angle compared to cup #1 and cup #2. All cups exhibited contact angles of ∼100°. However, dipping the cups in silicone oil followed by air drying significantly lowered the contact angle to ∼74° with a statistically significant reduction for cup #3.

**Figure 2.**
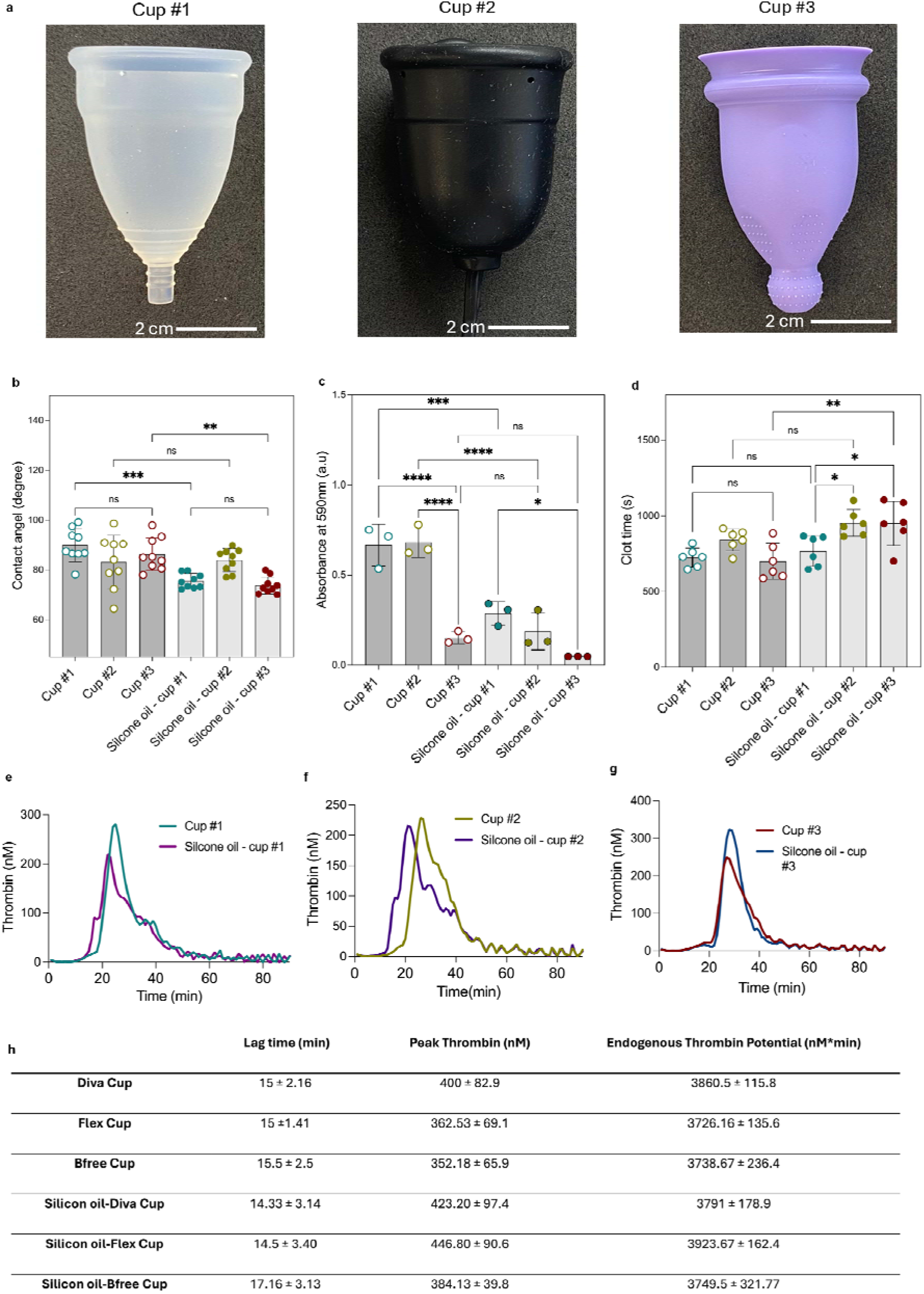
Structural characteristics of different menstrual cups. (a) Depicts the different commercial menstrual cups used for this study. (b) Contact angle of whole human blood on menstrual cups. (c) Biofilm formation of *E. coli* on the cup surface, as determined by the crystal violet method. (d) Clotting time of recalcified citrated human plasma on the cup surface. (e-g) Thrombograms of commercial menstrual cup. (h) Thrombogram-derived parameters, including lag time, peak thrombin, and endogenous thrombin potential. Significant levels are defined as follows: not significant (ns), p-value > 0.05; significant (*), p-value < 0.05; highly significant (**), p-value < 0.01; and very significant (***), p-value < 0.001.

In terms of resilience to biofouling, in the absence of surface treatment with silicon oil, cup #3 showed a 3-fold decrease in *Escherichia coli* biofilm formation compared to the other two cups. Surface treating all three cups with silicone oil significantly lowered biomass adhesion to cups #1 and #2. In contrast, cup #3 showed no significant reduction in adhered biomass compared to the non-surface treated cup (**Figure 2c**). Evidently, the oil infusion during manufacturing of cup #3 is sufficient to repel biomass adhesion. Surface treatment significantly increased the clotting time for cup #3 (from 699 to 949 seconds) with no effect on the other two cups (**Figure 2d**). Meanwhile, the thrombogenicity of all cups was not affected by silicone oil infusion, as all six thrombograms remained consistent (**Figure 2e-g**). Lag times, peak thrombin, and endogenous thrombin potential were also consistent and nonsignificant between all three cups, with and without silicone oil (**Figure 2h and Figure S1-4)**. Therefore, all three cups met the same standard of thrombogenicity, rendering them safe for intravaginal use. Notably, from qualitative observations of the cups before and after silicone oil infusion, we noticed that upon 24-hour incubation with silicon oil, cups #1 and #2 swelled and increased in size, which raises concern for long-term use. Based on the characterization data, we selected cup #3, which was surface treated with silicon oil, for further development **(Figure S5)**.

### Chemical characterization and swelling of fibers

The Ca^2+^-crosslinked alginate fibers were produced using an in-house wet spinning setup by injecting alginate solution into a CaCl_2_ coagulation bath. Fibers were prepared with 1 or 2% alginate and CaCl_2_ concentrations of 1, 2, or 3%. FTIR spectra of the alginate powder and alginate fibers crosslinked with different concentrations of Ca^2+^ in the range of 400-4000 cm^-1^ showed successful crosslinking of the alginate fibers, with characteristic peaks present in all six formulations (**Figure 3a**). The peaks, and -CO bonds observed at 3260, 2950, and 1025 cm-1, respectively, and are associated with the stretching vibrations of -OH, -CH, and -CO bonds.^18,37–39^ Several weaker peaks are observed in the 900-980 cm^-1^ range, which was attributed to mannuronic acid. Furthermore, the broad peak in the range of 1590-1600 cm^−1^ and the smaller peak in the range of 1450-1490 cm^−1^ were attributed to the asymmetric and symmetric stretching vibration of the carboxylate group, respectively. ^18,37–39^ As illustrated by the FTIR spectrum, the calcium alginate fibers exhibited a sharper -COOH peak at 1590-1600 cm^−1^ compared to alginate powder due to electrostatic interaction between Ca^2+^ and the carboxyl group, further confirming successful crosslinking of alginate.^18^

**Figure 3.**
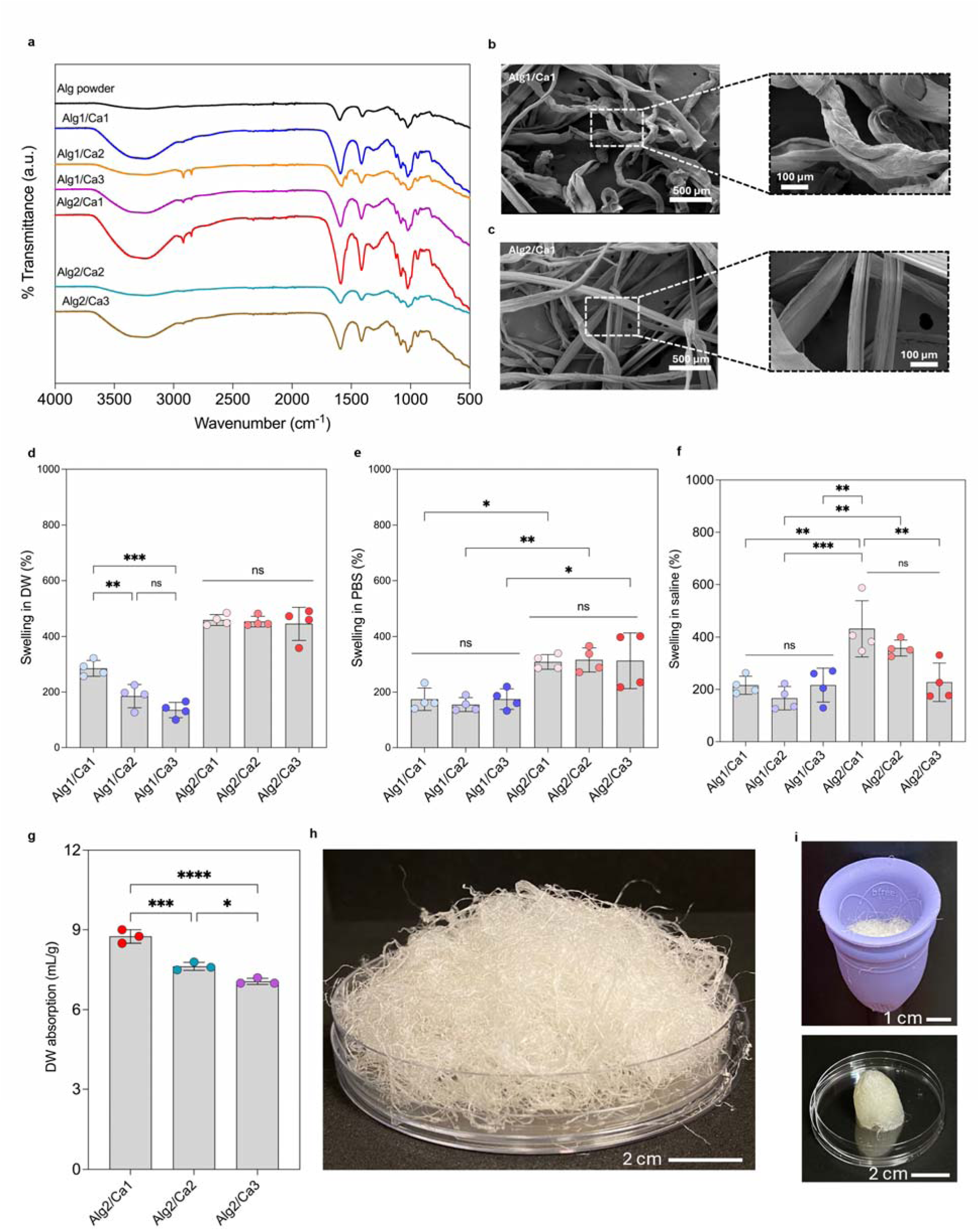
(a) FTIR spectrum of alginate powder in comparison to calcium-crosslinked alginate fibers at different concentrations of alginate and Ca^2+^ ions. SEM image of the (b) Alg1/Ca1 and **c.** Alg2/Ca1 fibers demonstrating the effect of alginate concentration on the fiber microstructure and surface morphology. (d) Swelling in Deionized water (DW), (e) PBS, and (f) saline for all 6 formulations with varying concentrations of alginate and CaCl_2_ as a cross-linker. (g) DW absorption of 2% alginate fibers crosslinked with 1, 2, and 3% calcium. (h) representative image of the air-dried alginate fibers. (i) Visual representation of the sample inside a menstrual cup before and after DW absorption test. Significant levels are defined as follows: not significant (ns), p-value > 0.05; significant (*), p-value < 0.05; highly significant (**), p-value < 0.01; and very significant (***), p-value < 0.001.

To evaluate the surface morphology of the fibers, SEM images were obtained from fibers with different formulations. The change in CaCl_2_ concentration did not result in visible differences in surface morphology, whereas the change in alginate concentration clearly affected the fiber microstructure. The SEM images of 1% and 2% alginate crosslinked with 1% CaCl_2_ showed marked differences in their surface morphology (**Figure 3b and c**). As demonstrated in the SEM images, 1% alginate (Alg1/Ca1) resulted in a more wrinkled and uneven surface, while the 2% alginate (Alg2/Ca1), led to a smoother and more uniform surface morphology. At higher alginate concentrations, a greater number of carboxylate (COO) groups are available for cross-linking with calcium ions (Ca²), resulting in a stronger and denser network with improved mechanical stability and reduced surface irregularities. Furthermore, at higher alginate concentrations, the molecular density in the fibers is higher, which contributes to a more organized structure and smooth, wrinkle-free fibers after drying. ^27^

The swelling of the fibers was measured in deionized water (DW), PBS, and saline. As shown in **Figure 3d-f**, 2% alginate formulations exhibited higher overall swelling capacities compared to 1% alginate, possibly due to the higher concentration of hydrophilic groups, including carboxyl (-COOH) and hydroxyl (-OH). In general, the swelling behaviour of the alginate fibers was dependent on alginate and calcium concentrations, as well as the ionic strengths of the test liquid (ionic strength: PBS>Silane>DW). Based on our observations, changes in Ca^2+^ concentration from 1 to 3% did not have a significant effect on the swelling properties of the 2% alginate formulation in DW and PBS (**Figure 3d and e**). In comparison, 1% alginate formulations showed increased swelling in DW at lower Ca^2+^ concentrations and remained unaffected by changes in Ca^2+^ in saline and PBS. Similar inconsistent traits were observed for the ionic strength effect of the test media. It is known that by increasing the ionic strengths of the media, the electrostatic repulsion of alginate increase due to the presence of cations like Na+, K+, and phosphate in PBS and Na+ in saline, respectively.^40^ As demonstrated in **Figure 3d-f**, Alg2/Ca1 exhibited a high swelling percentage in DW, saline, and PBS, with average swelling ratios of 458.03%, 431.08%, and 308.57%, respectively, due to its lower calcium crosslinking percentage and higher alginate concentration. Alg2/Ca2 also had high swelling ratios of 452.46%, 358.25%, and 315.80 % in DW, saline, and PBS, respectively. Moreover, Alg2/Ca3 showed the lowest swelling capacity of the 2% alginate samples (444.77%, 227.59%, 312.92% in DW, saline, and PBS, respectively). Regardless of the trends related to the effect of Ca^2+^ concentration and ionic strength of the media, Alg2/Ca1 exhibited the highest swelling percentage among the 2% samples in both DW and saline, while Alg2/Ca2 showed the highest swelling percentage in PBS. The water absorption capacity of the fiber was evaluated to determine its effectiveness in retaining fluids. DW absorption testing was conducted on samples with 2% alginate due to the higher concentration of hydrophilic groups and higher swelling ratio in 3 different media. As shown in **Figure 3g**, the absorption of Alg2/Ca1, Alg2/Ca2, and Alg2/Ca3 was measured to be 8.75, 7.63, and 7.06 mL of DW/g of fiber, respectively. In addition, the Alg2/Ca1 fiber a higher absorption capacity compared to the other samples due to its lower concentration of Ca2+, resulting in a lower hydrogel crosslinking density and higher absorption. In addition, **Figure 3h** shows a representative image of the visual appearance of air-dried alginate fibers (Alg2/Ca1), confirming the production of white, lightweight, and flexible thin fibers. This is followed by placing the fibers in a cup, as shown in **Figure 3i**, which depicts their appearance after water absorption. Based on swelling and liquid absorption performance, Alg2/Ca1 formulation was chosen for all subsequent experiments.

### Interaction of fibers with blood and bacteria

To create a proof of concept for commercial applications, the alginate fibers were die cut into tear-drop shapes (**Figure 4a**) demonstrating the customizability of the fibers allows them to be pressed into the shape of menstrual cups, thereby optimizing space. This also allowed us to conduct various blood and bacterial biocompatibility tests. For this objective, human citrated plasma clotting time analysis of 2% alginate fibers crosslinked with varying concentrations of Ca^2+^ showed a significant reduction in clotting time compared to the control (**Figure 4b**). Among the three formulations, Alg2/Ca1 had the shortest time to half-maximal absorbance 945.2 seconds (fastest clotting), most likely because the lower calcium concentration led to a reduced cross-linking density of the fiber and higher surface area, allowing for greater interactions with plasma proteins.^41^ To demonstrate the effect of crosslinking on the clotting time, tests using Factor XII deficient rabbit plasma were performed. This plasma was selected because without Factor XII, surface-induced contact activation cannot occur. The clotting assay results for control and factor XII-deficient rabbit plasma (control) are presented in **Figure 4c and d**. While there is a minor difference in the clotting outcomes between human and rabbit plasma—likely due to variations in plasma turbidity—Alg2/Ca1 exhibited faster clotting, whereas Alg2/Ca3 showed slower clotting. The blood coagulation cascade comprises two primary pathways: the intrinsic and extrinsic pathways, which culminate in the formation of a fibrin mesh that reinforces the platelet plug. The intrinsic pathway is initiated by Factor XII, which is activated by negatively charged surfaces. The activated form of factor XII, XIIa, activated factor XI. Subsequent steps of the intrinsic pathway require calcium ions, which act as crucial cofactors and facilitate the assembly of enzyme complexes. This includes the conversion of factor IX to factor IXa and the stabilization of factor IXa-factor VIII and factor Xa-factor V complexes^42^. As such, Ca^2+^ ions in calcium alginate fibers can promote blood clot formation and increase blood absorption. Moreover, the calcium ion concentration influences the porosity and surface area of alginate fibers, which plays a vital role in the intrinsic pathway. Our data indicate no difference in clotting time in factor XII-deficient plasma, suggesting that the variation in clotting times among the fibers is primarily due to the degree of crosslinking (**Figure 4d**).

**Figure 4.**
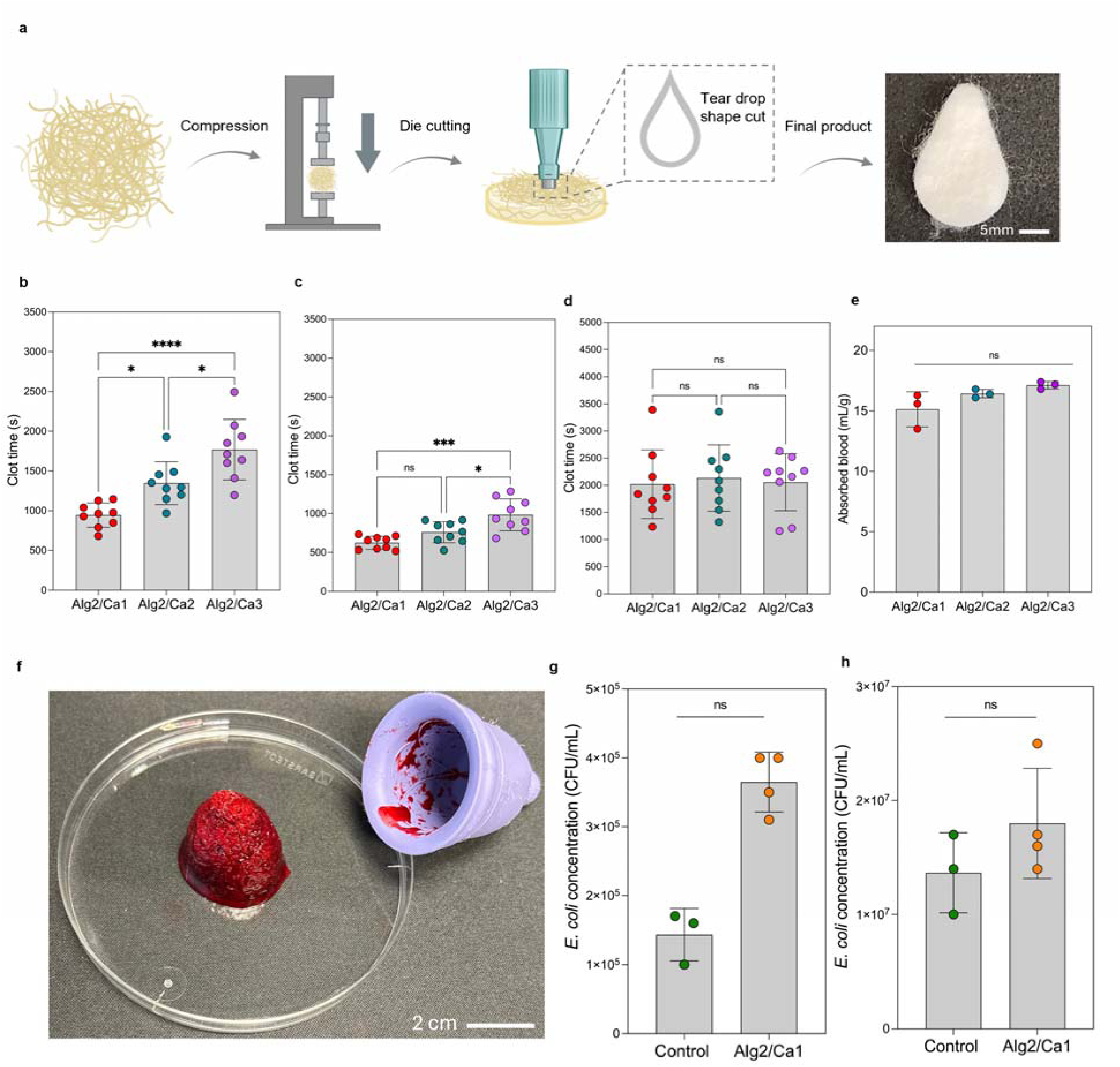
(a) depicts the compression and die cutting of the fiber to final product. Clotting ability of the fiber with the (b) Human citrated plasma **c.** rabbit citrate plasma and (d) factor XII knocked down rabbit plasma (e) Blood absorption of 2% alginate crosslinked with 1, 2, and 3% calcium (f) visual representation of the sample inside a menstrual cup before and after blood absorption test. Bacterial growth (CFU/mL) of *E. coli* incubated with alginate fiber after 7hrs incubation at 37°C with initial concentration of (g) 10^5^ CFU/mL and (h) 10^7^ CFU/mL. Significant levels are defined as follows: not significant (ns), p-value > 0.05; significant (*), p-value < 0.05; highly significant (**), p-value < 0.01; and very significant (***), p-value < 0.001.

In terms of blood absorption, shown in **Figure 4e**, there was no significant difference among 2% alginate samples crosslinked with different amounts of calcium.

The fibers showed higher absorption capacity in blood compared to deionized water (DW) due to the presence of Ca², which initiates blood clot formation. Although Alg2/Ca1 had a higher swelling ratio, Alg2/Ca3 had the highest blood absorption owing to the higher Ca^2+^ content, which resulted in increased blood clotting and higher absorption. This blood absorption ability is important for proper hygiene and convenient and mess-free removal of the menstrual cup as shown in **Figure 4f**.

Menstrual products and biomaterials with high absorption capacities, such as tampons and CMC, have been linked with the overgrowth of harmful bacteria, which can cause TSS through the production of bacterial toxins.^43^ Therefore, the interaction of the tablet with bacteria is an important design aspect that must be explored. To evaluate this, we incubated the alginate fibers with *E. coli* at high and low seeding densities for 7 hours—the average time the cup is worn before removal—and quantified the number of bacteria in the medium. Our results (**Figure 4g and h**) indicated that the Alg2/Ca1 fiber exhibited the highest absorption in water and showed no significant difference from other compositions in blood and had no significant impact on bacterial growth as illustrated by similar numbers of bacteria with and without fibers. Additionally, these results alleviate the concern that alginate, a polysaccharide, may serve as a nutrient source for microbes, thereby contributing to their proliferation. The results of these tests indicate that the calcium alginate fibers do not inhibit or promote additional growth of bacteria. As such, they are believed to be safe for consumer use in menstrual cups.

## Conclusion

In summary, we report a plant-based absorbent fibrous material designed to address one of the primary limitations of the menstrual cup, which is concerns about hygienic use. Incorporating our alginate-based fibers with commercially available menstrual cup enabled absorption of 15 mL/g of blood (a 1gr tablet fits in a standard 15mL cup). This fibrous tablet significantly improved the practicality and user friendliness of menstrual cups, making them comparable to pads and tampons in terms of absorption and user compliance in the market. Moreover, silicone oil surface treatment of the menstrual cups significantly decreased biofouling. Our results suggest that incorporating superabsorbent fibers into silicone oil-infused cups can enhance cup hygiene and the user experience, potentially increasing the acceptance of menstrual cups as an eco-friendly, cost-effective alternative to traditional menstrual products. Nevertheless, further studies are necessary to explore the optimal manufacturing protocols for an assembly line to fabricate fibers with smaller diameters. Smaller fiber diameters can address challenges faced with packaging the fibers into tablet form, ensuring minimal material is needed to achieve the necessary blood absorption levels of commercial menstrual cups. Additionally, conducting tests for flushability and biodegradation in soil, freshwater, and seawater as future steps of this work can revolutionize menstrual products, shifting current manufacturing best practices towards sustainable and environmentally friendly materials.

## Method and Experimental Section

### Material

Sodium alginate (SA, higher molecular weight) was purchased from Vyld company., Ltd. (berlin, Germany). Calcium Chloride Anhydrous (CaCl2) was purchased from Canada. Phosphate-buffered saline (PBS) tablets were purchased from VWR (Mississauga, ON, Canada). Silicone oil and silicon-based Cup (cup #3) were supplied by Bfree Cup company (Hamilton, ON, Canada). Diva (cup #1) and Flex Cups (cup #2) were purchased from Shoppers (Hamilton, ON, Canada) Bacterial stocks of *Escherichia coli* BL21 and Dh5a, were obtained from McMaster university (ON, Canada). Pooled citrated human plasma, rabbit factor XII-knocked down plasma, rabbit citrated plasma, and human whole blood were collected from healthy donors as previously described. ^44,45^ Venous blood was collected in tubes containing 3.2% v/v sodium citrate from healthy volunteers. A signed written consent was collected from donors and all procedures were approved by the McMaster University Research Ethics Board. Glacial Acetic acid and Crystal violate were purchased from Fisher chemical.

### Preparation of Sodium alginate Fibers

In this study, the technique of wet spinning was leveraged to fabricate sodium alginate (SA) hydrogel fibers crosslinked with Calcium ions by injecting SA solutions into a calcium chloride (CaCl_2_) coagulation bath. In detail, 1 and 2%w/v of SA solution was prepared by dissolving SA in distilled water while stirring at 400 rpm for 5 hours at 50 °C. The SA solution was extruded into a coagulation bath at varying concentrations of CaCl_2_ using a 25-gauge needle and a syringe pump at a flow rate of 3mL/min (**Table 1**). Afterwards, the fibers were washed 3 times with distilled water and dried at ambient temperature for 36 hours to obtain superabsorbent SA fibers. The preparation process of the superabsorbent SA fibers is shown in Fig.1.

**Table1.** The formulation of synthesized SA Fibers.

| Sample | $\text{CaCl}_2$ (% w/v) | Alginate (% w/w) |
| --- | --- | --- |
| Alg1/Ca1 | 1 | 1 |
| Alg1/Ca2 | 2 |  |
| Alg1/Ca3 | 3 |  |
| Alg2/Ca1 | 1 | 2 |
| Alg2/Ca2 | 2 |  |
| Alg2/Ca3 | 3 |  |

### Scanning Electron Microscopy (SEM)

The air-dried samples were mounted on SEM stubs using a double-sided carbon tape were and coated with a 20 nm gold layer using a Polaron Model E5100 sputter coater (Polaron Equipment Ltd., UK). SEM images were acquired using JEOL 6610LV SEM at the Canadian Centre for Electron Microscopy (CCEM), McMaster University (ON, CA).

### Fourier Transform Infrared Spectroscopy (FTIR)

FTIR was conducted to confirm the crosslinking of SA with calcium ions in all formulations. FTIR spectra were obtained from SA powder and SA crosslinked with different concentration of calcium in the range of 4000-400 cm^−1^, at 2 cm^−1^ resolution with 32 scans (Bruker instrument, model Vertex70).

### Swelling Property and Absorption Test

The swelling properties of the dried fiber were determined by measuring the weight of the fibers before and after testing. The fibers were weighed, placed into a cell strainer, and soaked in distilled water (DW), phosphate-buffered saline (PBS) or saline. The cell strainer was then removed after 1 hour and gently wiped to remove the excess unabsorbed liquid. The weight of swollen fibers was measured, and the swelling ratio was calculated using the following equation:

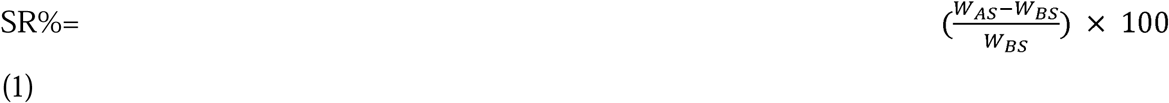

Where SR is defined as the mass of the absorbed fluid per mass of dried fiber, W_AS_ as the weight of the fibers after swelling and W_BS_ as the initial weight of dry fibers.

To measure the absorption capacities of the fibers, two different approaches were implemented. First, the immediate absorption of the fibers was assessed by dropwise addition of distilled water to the fibers followed by measuring the volume of absorbed. Next, the long-term absorption capacities were examined by first placing 1g of the fibers into menstrual cups and dispensing 25 mL of water into them over a time frame of 4–6-hours, using a syringe pump at a flowrate of 0.1 mL/min. At the endpoint, the fibers and remaining water in the cups were measured in order to assess the absorption capacities of the fibers. The values were reported as the volume of liquid absorbed per gram of fiber (mL/g).

### Thrombin generation Assay

The cups were cut into rings using biopsy punches with an inner diameter of 8 mm and an outer diameter of 10 mm. Afterward, 100 µL of plasma was added to each well of the plate and incubated for 10 minutes at 37°C. Then, 100 µL of the substrate mixture was added using a multichannel pipette. The substrate mixture contained 1 mM Z-GGR-AMC thrombin substrate (final concentration) and 15 mM CaCl (final concentration) in 20 mM HEPES. Fluorescence readings were taken for 90 minutes at 37°C and 1-minute intervals using the Technothrombin TGA protocol on a fluorescence plate reader (excitation: 360 nm, emission: 460 nm).

### Clotting Assay

The plasma clotting assay was conducted to investigate the clotting ability of the fibers as an essential feature that can improve their function as a menstrual fluid absorbent material. Fibers were weighed (15mg), rolled into a ring shape, and placed against the inner wall of the 96 well-plates. Empty wells were used as control. Subsequently, 180μL of different type of plasma such as citrated human plasma, rabbit factor XII-knocked down plasma, rabbit citrated plasma was added to each well, and absorbance was measured using a spectrophotometer (THERMO max) in kinetic mode at 405 nm with 6 second intervals. Due to the high presence of calcium in the fibers, the clotting assay was conducted in the absence of extra calcium. Clotting time was determined as the time to half-maximal absorbance. For the cup clotting assay, the ring-shaped cup segment was placed in a 96-well plate following the thrombin generation assay. Then, 75 µL of 20 mM HEPES buffer (pH 7.4) and 100 µL of human citrated plasma were added to each well containing the cup segment. Afterward, the plate was incubated for 15 minutes at 37°C. Next, 25 µL of 160 mM CaCl was added, resulting in a final CaCl concentration of 20 mM in the well. The same settings used for the clotting assay of the fibers were applied for readings.

### Whole human blood Absorption

The whole blood absorption test was performed to assess and simulate the blood absorption capacity of the fibers. For this purpose, 0.1g of the fiber was placed in a 15mL falcon tube and 2000 mL of citrated whole human was added gently. As mentioned earlier, due to the presence of Ca² ions in the fibers, no additional Ca² was added to initiate clotting. Excess blood was carefully removed after 30 minutes, and samples were weighed. The volume of absorbed blood was then calculated based on the blood’s density (1060 kg/m³).

### Bacterial Growth Assay

Frozen bacterial stocks were stored at −80°C in 25% v/v glycerol. Overnight bacterial cultures were prepared by inoculating 10 mL of bacterial media with glycerol stock. *E. coli* BL21 strains were cultured in LB media. The inoculated media was incubated at 37 °C and 180 rpm for 16–18 h to promote bacterial growth. Subsequently, the bacterial cultures were centrifuged at 7000 rcf for 15 minutes to retrieve bacterial pellets, the supernatant was decanted, and the pellets were washed with PBS 3 times. The undiluted concentration of the bacteria was 10^9^ CFU/mL. To determine the effect of the calcium alginate fibers on bacterial growth, both spot tests and tear drop tests were conducted. For the spot test, bacterial solutions with a concentration of 10^9^ CFU/mL were diluted to 10^7^ and 10^5^ CFU/mL. Then, 300 µL of this solution was added to 0.006 g of sterile fiber and incubated for 7 hr. Afterward, the concentration of the bacteria was determined using the tear drop and spot tests which involved plating the bacterial solutions on MacConkey agar and incubating overnight at 37°C.

### Crystal violates (CV) assay

The cups were cut into disk shapes using 10 mm biopsy punches and placed in a 48-well plate. An overnight culture of *E. coli* (DH5α) with an OD of 1 was diluted 1:40 in LB media. Then, 600 µL of the diluted bacterial culture was added to each well and incubated at 37°C for 24 hours. After incubation, the bacterial suspension was removed, and the cups were washed three times with sterile PBS to remove excess bacteria. The cups were then submerged in 500 µL of 0.05% crystal violet (CV) solution for 10 minutes. Next, the cups were rinsed five times with PBS to remove excess stain and left to dry overnight. The stained cups were then placed in 500 µL of 30% acetic acid to dissolve the bound CV dye, followed by brief vertexing. Finally, 100 µL of the CV-acetic acid solution was transferred into a 96-well plate, and the absorbance was measured at 590 nm using a Synergy Neo2 BioTek plate reader.

### Statistical analysis

All results are presented as mean ± standard deviation (SD). Unless otherwise stated, experiments were performed at least 3 times. Significance of differences between groups was determined using one or two-way analysis of variance (ANOVA) followed by GraphPad Prism 10 (GraphPad Software, San Diago, CA) were used to create graphs. For all analyses, P < 0.05 was considered statistically significant.

## Data Availability

All data produced in the present study are available upon reasonable request to the authors

## Acknowledgements.

T.F.D. and Z.H. acknowledge the support from the Canada Research Chairs Program. T.F.D. and Z.H. also acknowledge the support from Natural Sciences and Engineering Research Council (NSERC) of Canada through the Canada Discovery Grant and the Ontario Early Researcher Award.

## Supporting Information Available

thrombogram parameters—such as lag time, peak thrombin, time to peak thrombin, and the area under the thrombogram curve—as well as visual representations of the swelling behavior of cups #1 and #2 after incubation with silicone oil.

