## Supporting information for "Self-cleaning menstrual cups with plant-based biodegradable superabsorbent fibrous tablets for hygienic and sustainable Period Care"

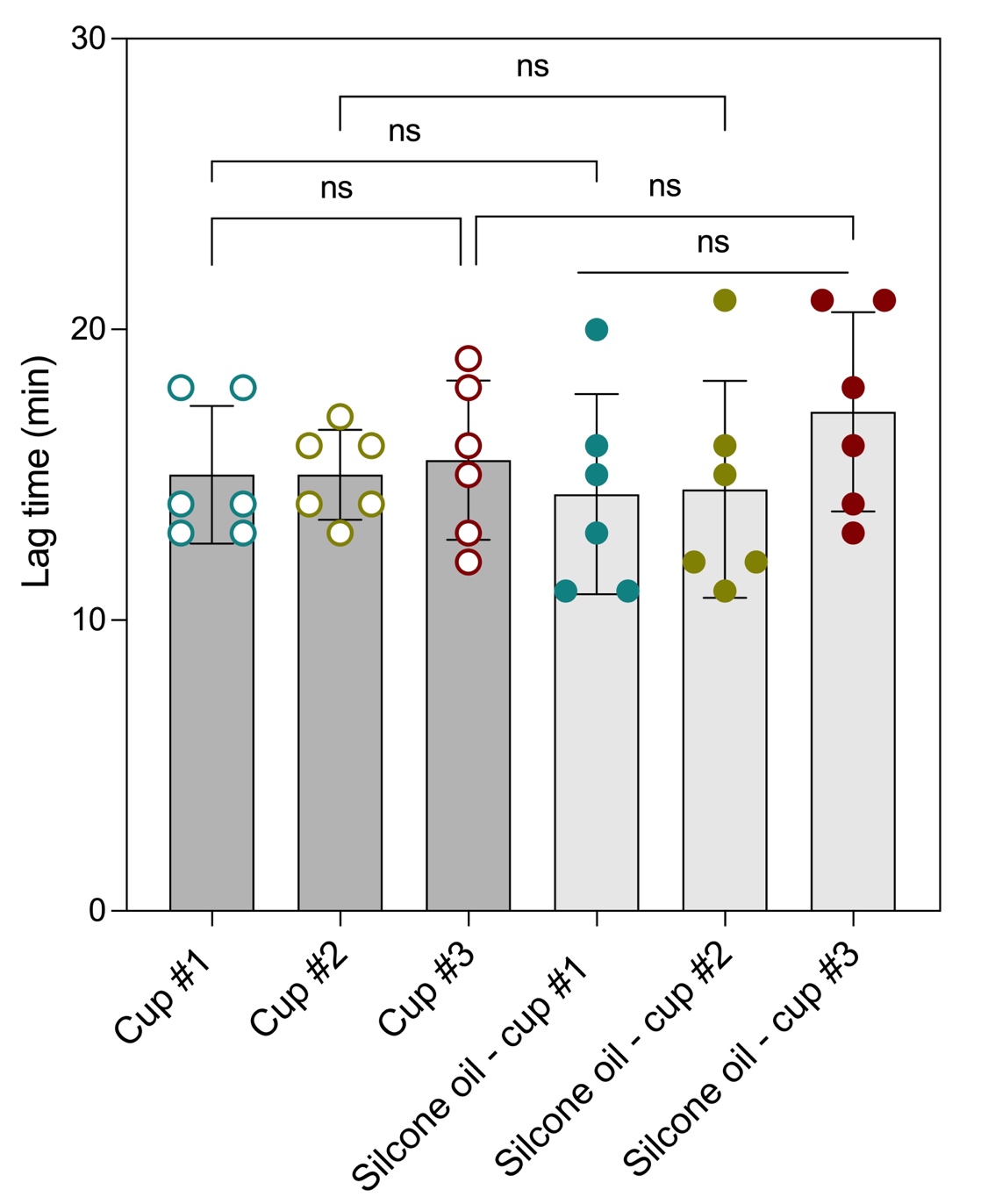

**Figure S1. Illustration of** lag time in the thrombogram. The lag time represents the interval between the start of the assay and the initiation of thrombin generation, reflecting the delay before measurable thrombin activity begins.

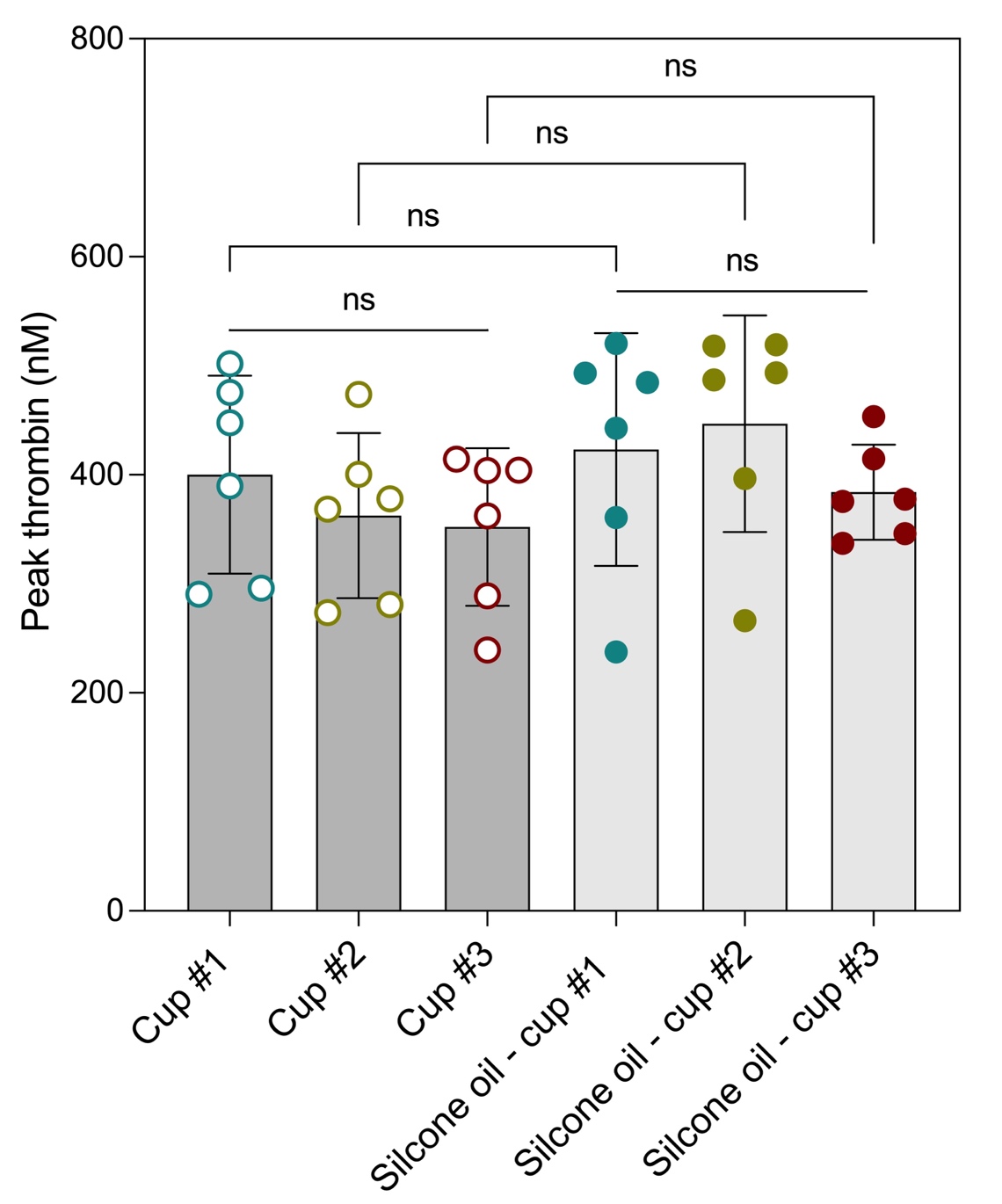

**Figure S2. Illustration of** peak thrombin in the thrombogram. The peak thrombin value indicates the maximum concentration of thrombin generated during the assay.

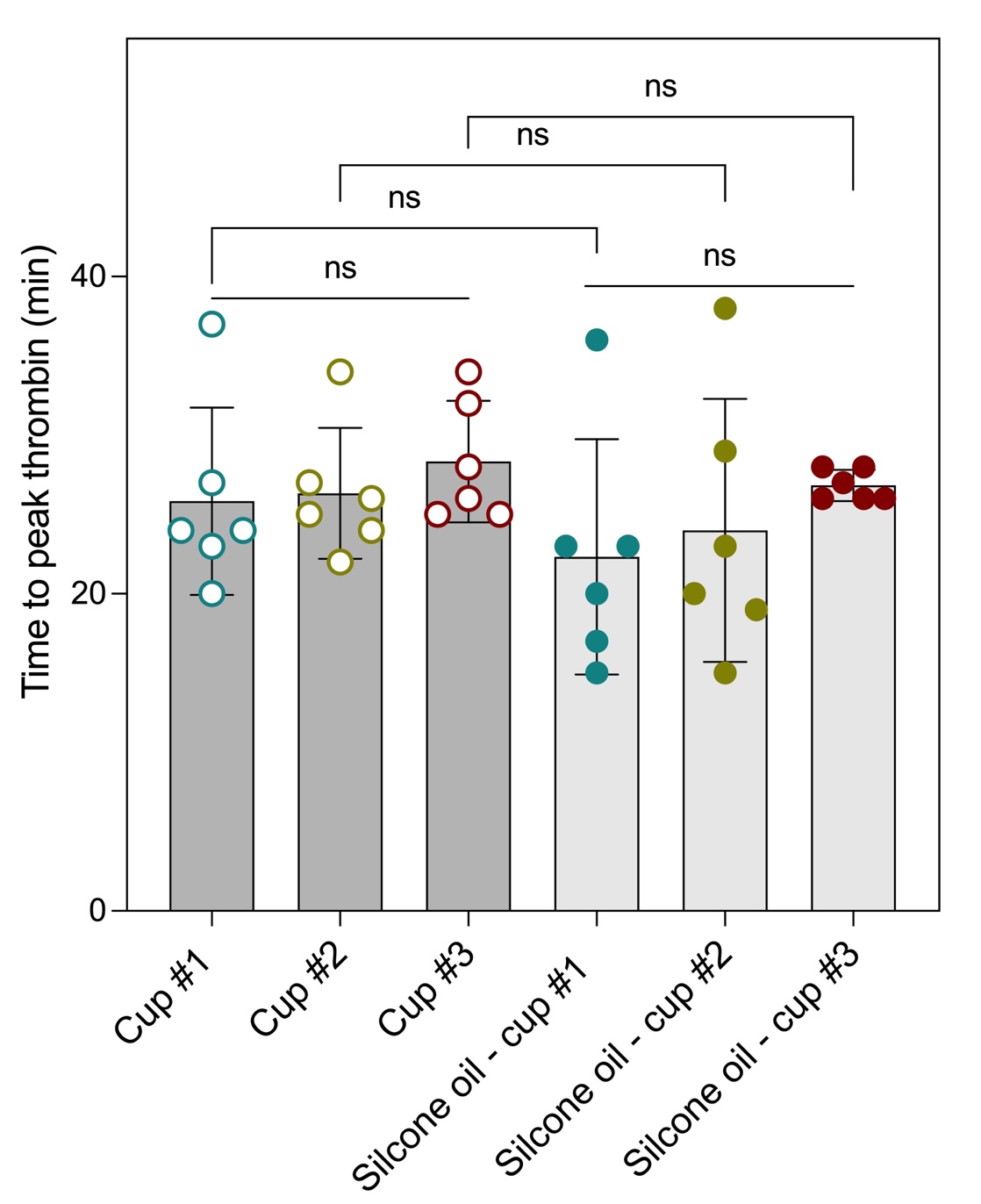

**Figure S3.** **Illustration of** time to peak thrombin in the thrombogram. This parameter denotes the duration between the initiation of the assay and the point at which the maximum thrombin concentration is reached.

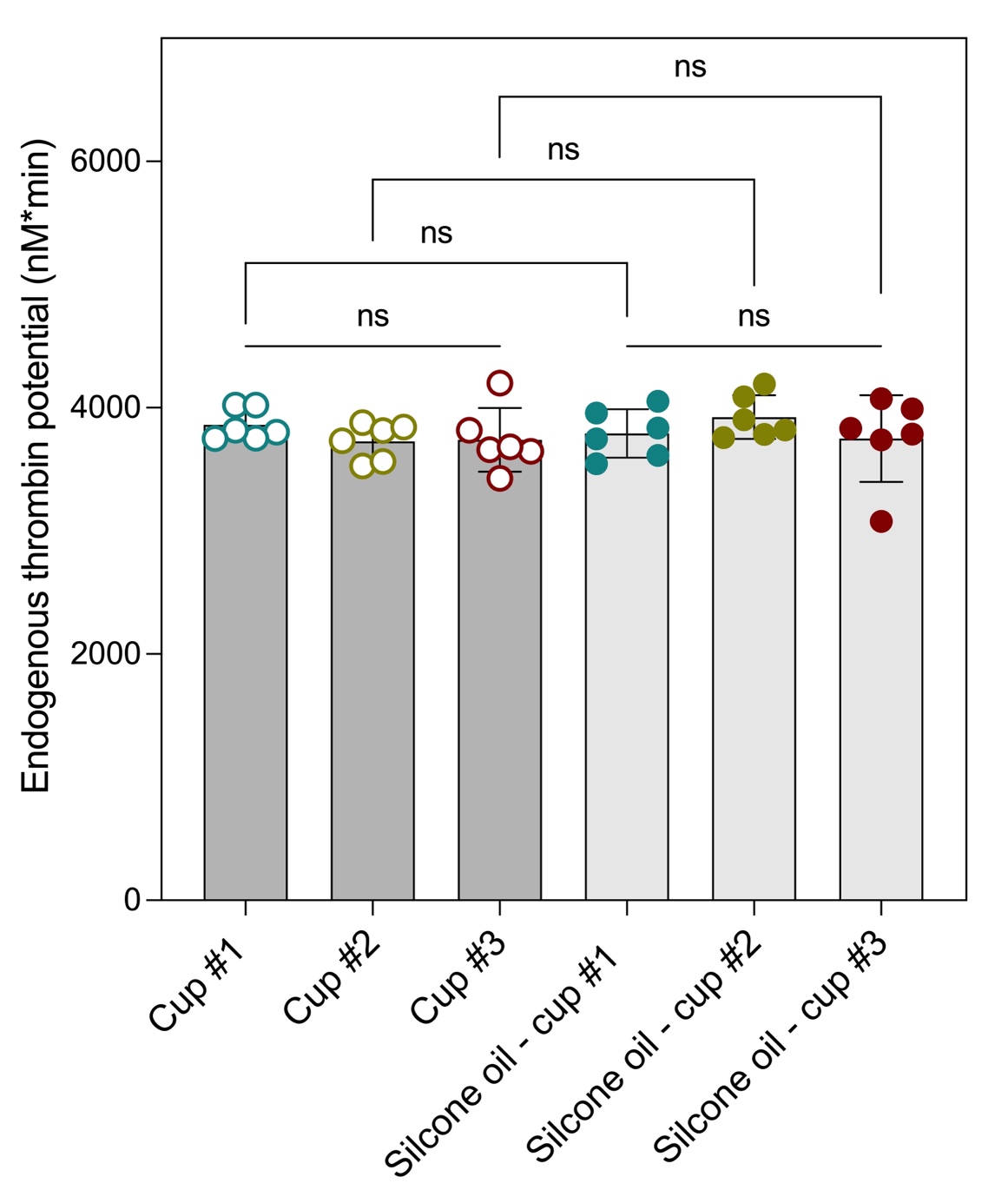

**Figure S4.** **Illustration of** endogenous thrombin potential (ETP) in the thrombogram. ETP represents the area under the thrombin generation curve.

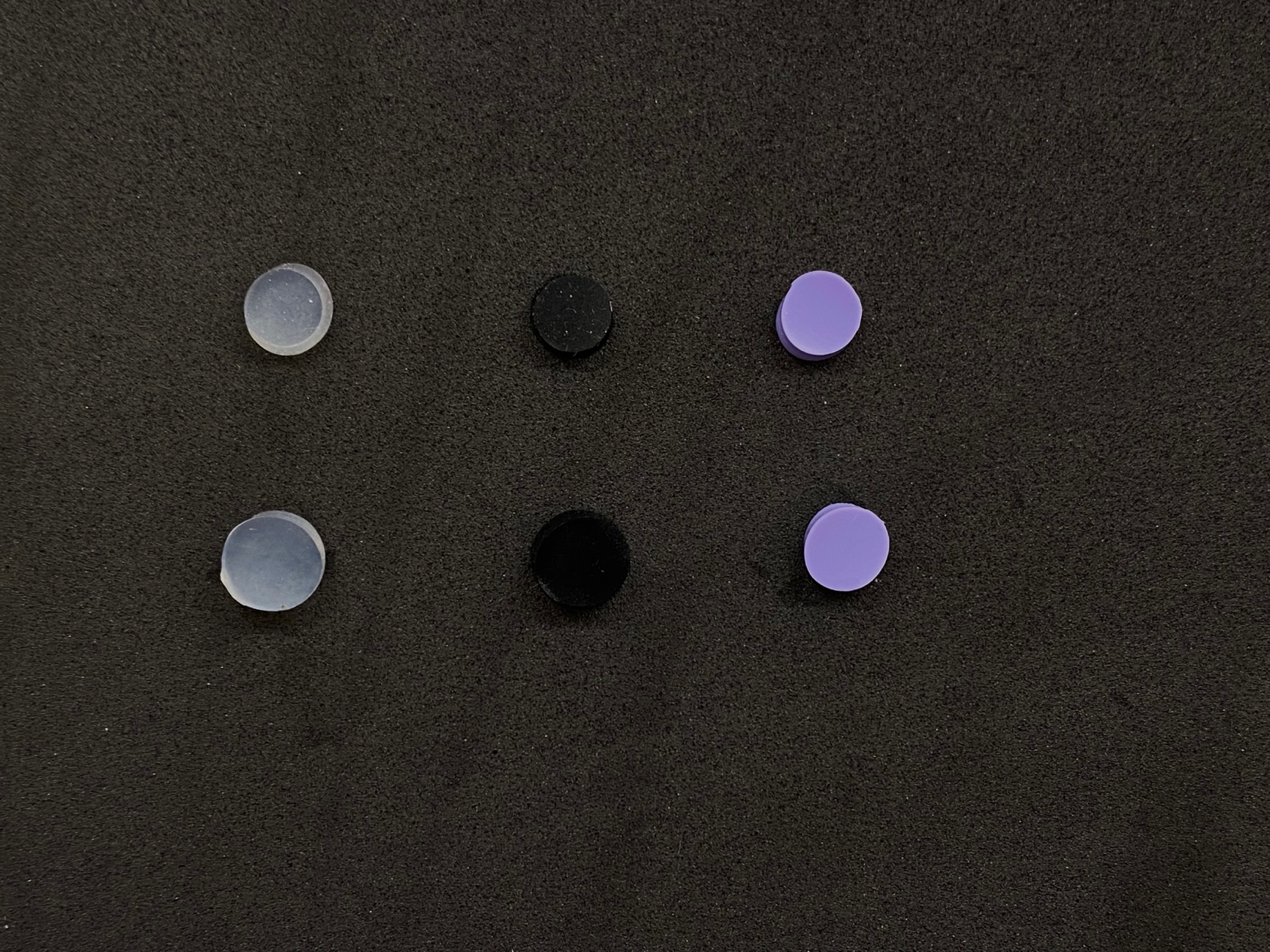

Cup #1

Cup #2

Cup #3

8mm

**Figure S5**.depict the swelling of cup #1 and 2 after 24hr incubation with the silicon-oil.
